# Multisystem Complications of Monkeypox: A Systematic Review from Low- and Middle-Income Countries in Africa

**DOI:** 10.1101/2025.03.19.25324244

**Authors:** Alex Mwangi Kihunyu, Abdulnaim Mohamed Hussein, Margaret Murugi Kioni, Lucy Ogoi, Sherlyn Chemutai, Jerome Kiungu, Abdalla Mwita, Rebecca Muiruri Nyakeru, Vernon Ipomai, Stacy. O. Akinyi

## Abstract

**Introduction:** Monkeypox, a zoonotic viral disease primarily endemic to Africa, has seen a global resurgence, raising concerns due to its potential for severe multisystem complications. While often self-limiting, the burden of monkeypox is heightened in low- and middle-income countries (LMICs), particularly in Africa, where healthcare resources are limited.

**Objective:** To review the multisystem complications of monkeypox in LMICs across Africa, highlighting clinical challenges and gaps in healthcare management.

**Methods:** A comprehensive systematic review was conducted using PubMed and Google Scholar to identify studies addressing monkeypox complications. Relevant literature covering dermatological, respiratory, neurological, gastrointestinal, cardiovascular, and ocular systems was reviewed, with a focus on clinical complications and healthcare constraints in African LMICs.

**Results:** The review identified significant multisystem complications of monkeypox infection, including impetigo, cellulitis, and septic shock in the dermatologic system; bronchitis and severe pneumonia in the respiratory system; encephalitis and seizures in the neurologic system; haemorrhagic colitis, gastrointestinal bleeding, and severe dehydration in the gastrointestinal system; and viral myocarditis, pericarditis, congestive heart failure, arrhythmias, thrombotic events, and dilated cardiomyopathy in the cardiovascular system. In African LMICs, these complications are exacerbated by limited diagnostic tools, antiviral therapies, and inadequate healthcare infrastructure. Socioeconomic challenges, including poor healthcare access and limited public health awareness, further compound the weaknesses in the health system.

**Conclusion:** Monkeypox presents complications across multiple organ systems. In African LMICs, healthcare barriers exacerbate disease burden. Strengthening infrastructure, improving diagnostics, and increasing awareness are key to mitigating the impact of monkeypox complications.

## 1.0 Introduction

Monkeypox is an emerging zoonotic disease caused by the monkeypox virus, an orthopoxvirus that has remained endemic in Central and Western Africa for over five decades (1). The disease is particularly prevalent in 11 countries within these regions, with thousands of cases reported annually (2). The virus is suspected to have reservoir hosts among African rodents and small mammals, such as the African striped squirrel (3). Although most cases of monkeypox are self-limiting, the case fatality rate ranges from 1% to 10%, depending on the viral clade, with the highest mortality seen in children across Africa (4). While monkeypox shares clinical similarities with smallpox, it differs in its zoonotic origins and generally presents with a milder course (5).

Monkeypox is well-documented for its acute phase symptoms, including fever, headaches, muscle aches, back pain, low energy, and rashes lasting 2–4 weeks. Swollen lymph nodes are also common (6). In monkeypox, skin lesions, particularly in the anogenital region, are a characteristic manifestation, with the median time from the onset of lesions to dry crust formation being around 10 days. Other symptoms include proctitis, tonsillitis, penile edema, abscesses, and exanthems, with rectal pain being significant (7;8). Newborns, children, pregnant individuals, and people with immune deficiencies, such as those with advanced HIV, are at higher risk of severe disease and death (6).

A recent study in the United Kingdom indicated that severe complications, such as hemorrhagic pustules and secondary bacterial infections, are more common in African patients. Among these patients, 64% require hospitalization, compared to European patients, who experience less severe symptoms, with approximately 10% being hospitalized (9). Additionally, reported complications in Africa include pitted and deforming scars, hyperpigmentation, secondary bacterial infections, bronchopneumonia, keratitis, corneal ulceration, blindness, sepsis, dehydration, vomiting, diarrhea, and encephalitis (10,11). A recent study from Nigeria demonstrated that the monkeypox virus can invade the central nervous system, leading to neurological complications such as chronic headaches, seizures, and photophobia (12). Furthermore, anorectal pain due to lesions, as well as complications like myocarditis and epiglottitis, have been identified (13).

This study aims to review the multisystem complications of monkeypox infection across diverse patient populations in Africa. It seeks to identify variations in disease progression, recovery, and long-term effects among different demographic groups, including children, pregnant individuals, and those with underlying health conditions, to enhance long-term care and improve patient outcomes.

## 2.0 Methods and Materials

A systematic review has been conducted following the Preferred Reporting items for SR and MA (PRISMA) 2020 guidelines ensuring transparency and rigor.

### Search strategy

A comprehensive literature search was conducted in two electronic databases, PubMed and Google Scholar. The search terms used included: “monkeypox,” “monkeypox and Africa,” “monkeypox” AND “complications,” “monkeypox” AND “long-term outcomes,” “monkeypox” AND “Central Nervous System in Africa,” “neurologic” AND “complications of monkeypox,” “dermatologic” AND “complications of monkeypox,” “respiratory system” AND “monkeypox in Africa,” “cardiovascular” AND “complications of monkeypox,” “musculoskeletal complications” AND “monkeypox,” “ocular complications of monkeypox in Africa,” and “gastrointestinal complications of monkeypox in Africa.”

The search was restricted to articles published in English but no limits were placed on the year of publication to ensure the inclusion of both recent data and novel studies reflecting the evolving epidemiology, clinical manifestations, and management approaches of monkeypox infection. Additionally, relevant studies were identified through a manual search of key references. The collected references were imported into Rayyan software for deduplication. Following this, the references were screened based on the established inclusion and exclusion criteria (Table 1).

**Table 1:** Inclusion and exclusion criteria.

| Criterion | included | Excluded |
| --- | --- | --- |
| Study design | Review articles, Case reports, Case Series, Observational studies | Randomized Control Trials, Quasi experiment studies, Opinion pieces, editorials, non-peer-reviewed articles |
| Population | Human participants diagnosed with monkeypox in low- and middle-income countries (LMICs) in Africa. | Animal studies, and studies focusing on non-African populations. |
| Outcomes of interest | Multisystem complications of monkeypox | Others diagnoses |
| Accessibility | Abstracts and full text accessible | Abstracts and full text non-accessible |
| Language | English | Non-English |

### Study selection process

Two reviewers independently conducted the study selection process using Rayyan software. Initially, all titles and abstracts were screened to exclude studies that did not meet the inclusion criteria. Full-text articles of the remaining studies were then retrieved and thoroughly reviewed to determine their final eligibility. In cases where disagreements arose between the two reviewers, conflicts were flagged for further discussion. The reviewers first attempted to resolve differences by discussing their reasoning. If consensus was not reached, a third impartial reviewer was consulted to facilitate the discussion. When necessary, the third reviewer made the final decision based on the inclusion and exclusion criteria and the study’s relevance. The study selection process was documented using the PRISMA flow diagram. (Figure 1)

**Figure 1:**
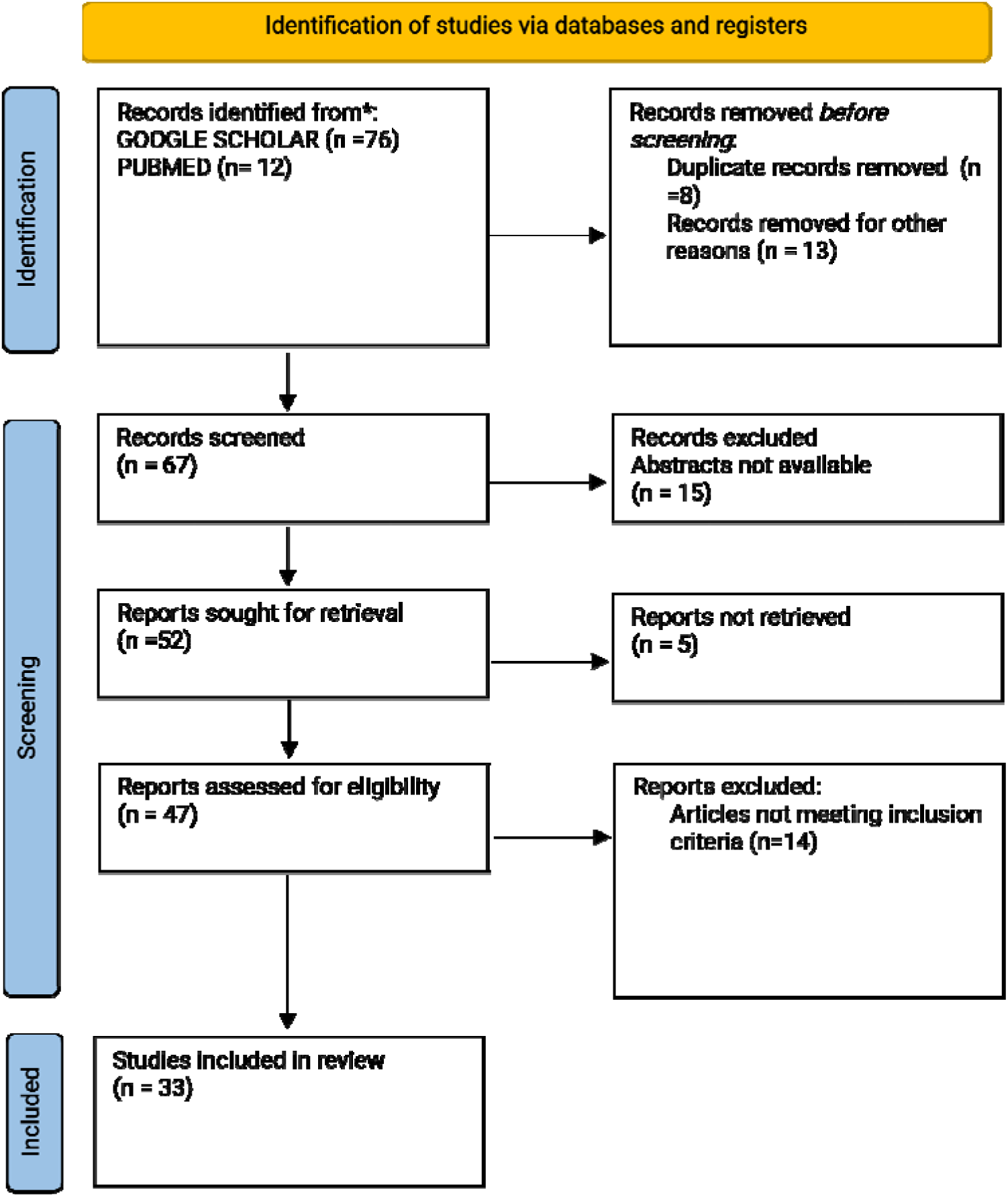
PRISMA flowchart

### Data Extraction

Data extraction was conducted using a standardized spreadsheet. Two independent reviewers extracted data from each included study, capturing key information such as sample size, patient demographics, affected organ system, prevalence/incidence of systemic complications, and morbidity and mortality rates. Any discrepancies in data extraction were resolved through discussion between the reviewers.

### Data Synthesis

Data synthesis was conducted systematically to assess the multisystem complications of monkeypox in low- and middle-income countries (LMICs) in Africa. Google Sheets and the X Miner extension were utilized to organize and summarize the extracted data, particularly focusing on age distribution, disease complications and morbidity and mortality data within the included studies.

The synthesis further examined the distribution of multisystem complications, highlighting trends in organ involvement, morbidity, and mortality rates. Additionally, variations in the incidence and prevalence of complications were explored across different patient demographics and healthcare settings. This structured approach provided a comprehensive understanding of the burden of monkeypox complications in African LMICs, offering insights that may inform targeted healthcare interventions.

## 3.0 Results

### Screening and Study Selection

A total of 88 records were identified through the initial search. After duplicate removal, the remaining studies were screened based on the inclusion criteria. Following a full-text review, 33 studies met the eligibility criteria and were included in the qualitative synthesis. A detailed PRISMA flowchart illustrating the selection process is provided in Figure 1.

### Baseline Characteristics

The study population encompassed individuals of all ages, reflecting the broad demographic impact of monkeypox. A comprehensive summary of the multisystemic complications associated with monkeypox is presented in Table 1.

### Quality Assessment

The quality of included studies was evaluated using the Newcastle-Ottawa Scale (NOS). Studies scoring ≥7 were classified as high quality, while those scoring <7 were considered low quality due to potential biases. Overall, nine studies were deemed high quality, whereas five studies exhibited limitations in cohort comparability, leading to a higher risk of bias.

### Outcomes

The key findings from the included studies, summarizing the clinical complications and outcomes of monkeypox across multiple organ systems, are presented in Table 1 and Figures 1, 2, and 3.

**Table 1.0:** Summary of Monkeypox-Associated Complications Across Organ Systems.

| Organ System | Key Findings | Geographic Distribution | Age & Sex Trends | Implications for LMICs |
| --- | --- | --- | --- | --- |
| **Dermatologic** | Skin lesions progress through macules, papules, vesicles, pustules, and scabs. Secondary bacterial infections (impetigo, cellulitis, sepsis) and post-inflammatory hyperpigmentation are common. Residual scarring is a significant burden. | High prevalence in the Democratic Republic of Congo (DRC), Nigeria, Ethiopia, Kenya, Mozambique, Rwanda, Uganda, and Zambia. | Median age shifted from 4 years (1970s) to 21 years (2010s). More common in unvaccinated individuals. | Limited antibiotic access, poor WaSH infrastructure, and inadequate wound care increase morbidity. |
| **Musculoskeletal** | Myalgia, arthralgia, and muscle weakness persist post-infection. Chronic joint pain and myositis reported. Cases of muscle deformities observed. | Reported in Nigeria, West Africa, and DRC. | No significant sex differences. Young and elderly at higher risk due to nutritional deficiencies. | Scarcity of physiotherapy and pain management services, along with malnutrition, worsens outcomes. |
| **Central Nervous System (CNS)** | Headache, myalgia, fatigue, confusion, encephalitis, seizures, and psychiatric symptoms. High mortality rates in severe cases. | DRC (2007-2011: 216 cases, 49 headaches, 15 myalgia, 5 visual deficits). Nigeria (40 cases: 19 headaches, 25 myalgia, 1 suicide). | Mean age: 27.8 years (range 28 days-43 years). Mortality rate: 29.4%. | Diagnostic challenges due to limited CSF analysis capabilities. High cost of vaccines limits prevention. |
| **Cardiovascular** | Rare but includes myocarditis and pericarditis. Viral myocarditis may progress to heart failure. | Few cases reported, primarily in Nigeria and DRC. | No significant data on age/sex distribution. | Limited diagnostic and long-term care facilities make early detection difficult. |
| **Ocular** | Eyelid and conjunctival involvement (conjunctivitis, ulcers, keratitis, uveitis). Rare cases of dacryocystitis and optic neuritis. | Reported in Nigeria (25% of patients) and DRC (23.1%). | No age/sex distinction provided. | Lack of ophthalmologic care increases risk of long-term visual impairment. |
| **Respiratory** | Bronchopneumonia, pleural effusion, and ARDS. Higher mortality rates in children and immunocompromised individuals. | High burden in DRC and Nigeria (15-20% of MPX patients develop respiratory complications). | 65% of severe cases under 15 years old. Slight male predominance (55-60%). | Poor access to critical care and ventilation support contributes to high mortality. |
| **Gastrointestinal** | Anorexia, diarrhea, nausea, vomiting, abdominal pain, hemorrhagic colitis, and gastrointestinal bleeding. | DRC (1996-1997: 11% of 419 cases had diarrhea). Recent hemorrhagic colitis cases in Nigeria. | 85% of affected cases under 16 years old. | Poor sanitation, dehydration, and lack of rehydration therapy exacerbate complications. |

**Figure 1.0:**
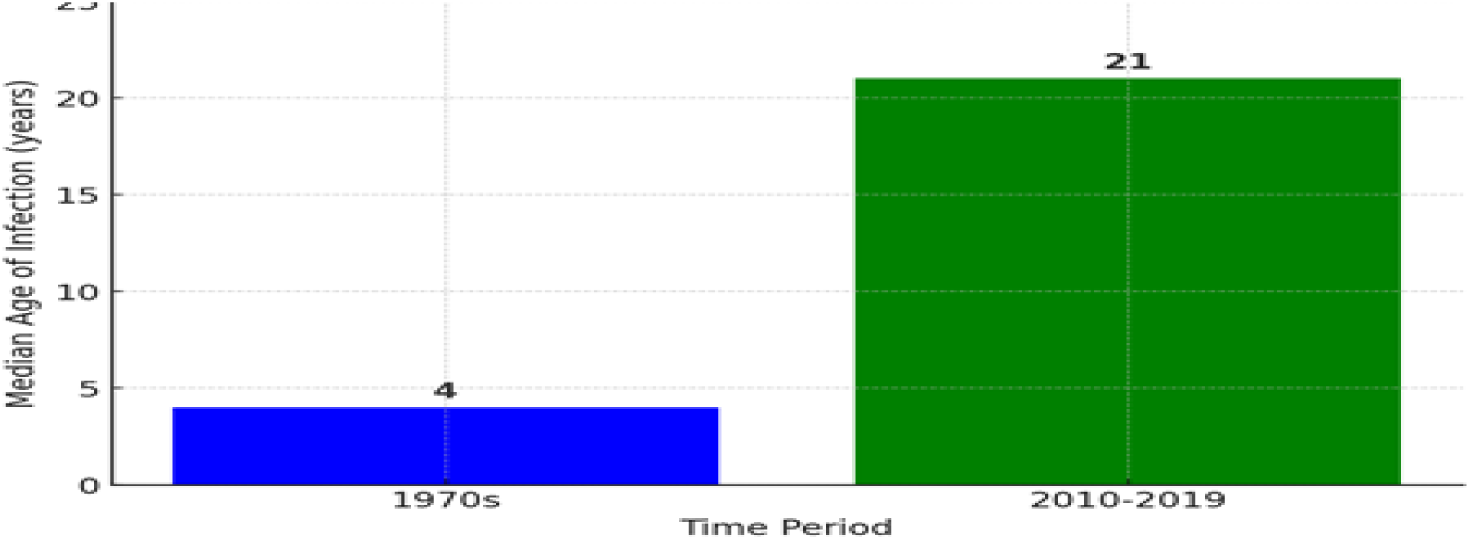
Shift in Monkeypox case distribution from children in the 1970s to young adults in recent years.

**Figure 2.0.**
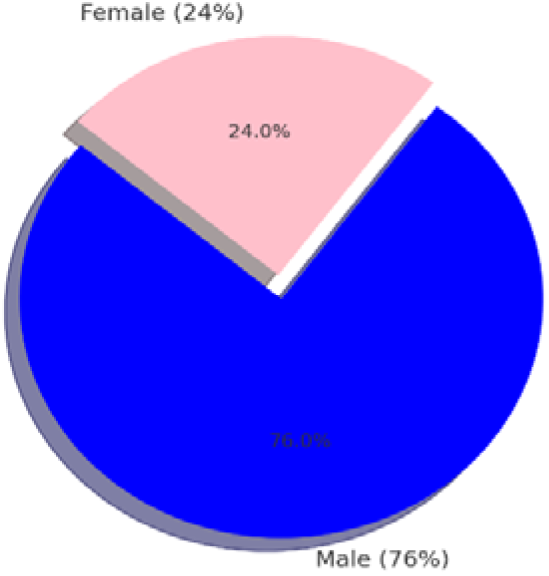
sex distribution of Mpox-related respiratory complications in Nigeria and the Democratic Republic of Congo.

**Figure 3.0:**
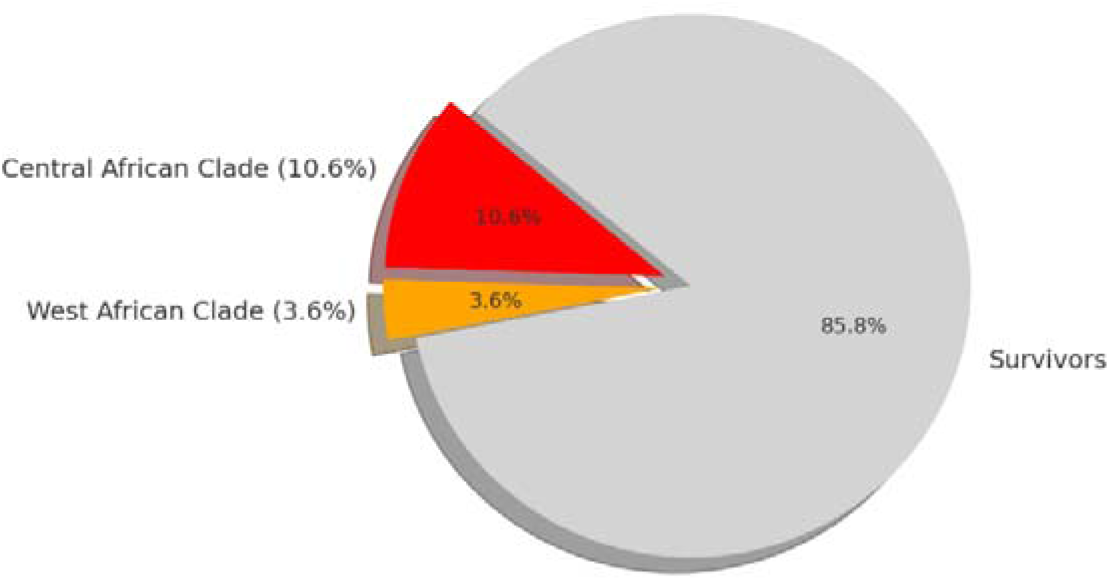
Case Fatality Rate (CFR) of different Mpox clades.

## 4.0 Discussion

### 4.1 Dermatologic System

One of the most distinctive and diagnostic features of monkeypox is its dermatological manifestations (14). The skin lesions progress through five stages: macules, papules, vesicles, pustules, and scabs (14). These lesions are most concentrated on the face and extremities, often following a centrifugal pattern that spreads toward the trunk, with severity and distribution influenced by geographic location and timeliness of medical intervention (15).

A systematic literature review for Mpox in humans from 1970 to 2019 reported a marked increase in cases with time, most notably in the Democratic Republic of the Congo (DRC). The literature revealed a shift in the median age of infection from young children (4 years) in the 1970s to young adults (21 years) in 2010–2019, suggesting a change in epidemiology possibly due to smallpox vaccination discontinuation (4).

Secondary bacterial infections are a common complication of Mpox skin lesions. Such infections lead to conditions such as impetigo and cellulitis, and in severe cases, sepsis (15). Limited access to antibiotics and unavailability of clean environments in rural African areas aggravate such complications, A study in rural South Africa revealed that community members often face challenges in sourcing antibiotics, leading to potential misuse and increased risk of complications (16). Additionally, assessments of healthcare facilities in countries like Ethiopia, Kenya, Mozambique, Rwanda, Uganda, and Zambia have identified deficiencies in water, sanitation, and hygiene (WaSH) infrastructure, further contributing to adverse health outcomes (17). Residual scarring and post-inflammatory hyperpigmentation (PIH) are serious long-term complications, predominantly in darker-skinned individuals (18). Such skin complications lead to social and psychological burdens, most significantly in populations with limited access to skincare and reconstructive surgery (19).

The Case Fatality Rate (CFR) varies in different clades of Mpox virus. For the Central African clade, a 10.6% CFR exists, and for the West African clade, a 3.6% CFR. The rates indicate how serious Mpox infections are and how skin complications should be treated in an attempt to reduce morbidity and mortality (4).

Early therapy, adequate wound management, and access to aseptic facilities are important in preventing complications from Mpox skin lesions. Such basic health services, however, in most African low- and middle-income countries (LMIC) are either absent or inaccessible significant drivers for Mpox transmission and severity (18). To address such challenges better, intensive research is required to delineate specific prevalence, incidence, and long-term outcomes of skin complications due to Mpox in diverse African LMIC populations. Such data are important for guiding focused interventions, optimal resource allocation, and enhancing overall patient outcomes in such environments.

### 4.2 Musculoskeletal System

Myalgia, arthralgia, and generalized weakness are frequently reported during the acute phase of monkeypox infection and often persist into the recovery phase (20). In an outbreak of monkeypox reported in Nigeria between 2017 and 2018, 63% of cases had myalgia (21). A 24 year old man unvaccinated from smallpox was found to have neck stiffness on presentation (22). These symptoms are believed to result from the body’s systemic inflammatory response to the virus, leading to muscle pain and joint inflammation (20). Many patients report muscle weakness and chronic joint pain lasting months after the acute infection has resolved, as observed in multiple cases (20). In addition, a case has been found to have deformities of facial muscles as a sequelae to healing of ulcerated facial lesions (23).

In LMICs, where rehabilitation services are scarce, individuals suffering from musculoskeletal complications face a higher risk of prolonged disability (24). Physical therapy and pain management services are often unavailable in rural areas, exacerbating the impact of reduced mobility and chronic pain (24). Nutritional deficiencies, common in LMICs, further contribute to these musculoskeletal issues, especially among children and the elderly (25). Recovery from viral-induced muscle and joint damage is slowed by deficiencies in calcium and vitamin D, both prevalent in food-insecure regions (25).

A case report from West Africa described a patient experiencing severe myositis post-recovery from monkeypox, which persisted for months and significantly limited daily activities (26). This underscores the need for comprehensive management strategies, including nutritional support and physiotherapy, to prevent long-term musculoskeletal disability in affected populations (26).

### 4.3 Central Nervous System

Neurological manifestations of monkeypox, including headaches, myalgia, malaise, fatigue, altered consciousness, agitation, anorexia, nausea, and vomiting, are increasingly recognized (27). Neuropsychiatric presentations such as confusion, encephalitis, seizures, and nonspecific symptoms like headaches and muscle pain have also been observed (28). Experimental data suggests that the monkeypox virus may access the central nervous system (CNS) via the olfactory epithelium and infected circulatory monocytes/macrophages (27). In Nigeria 40 confirmed monkey pox cases neurological complications identified included headache 19, myalgia 25, seizures 1, encephalitis 3, photophobia 9 anxiety and depression 11 and 1 suicide case. (29). Similarly in a DRC study between 2007 to 2011, out of 216 confirmed cases, neurological complications included headaches 49, myalgia 15, dizziness 3, visual deficits 5, confusion 4 and 11 fatigue cases. (29). A study by Shramana Deb et al, 2025, of the 18 cases, 12 were men (66.7%), four were women (22.2%), and two (11.1%) did not report age or sex. The mean (median) age was 27.8 (30.0) years, ranging from 28 days to 43 years. Of the said cases the outcomes were reported in 17 cases, 29.4% of them recovering fully, 41.2% recovering partially and 29.4% of them died. (30).

Diagnosing monkeypox involves clinical, epidemiological, and laboratory criteria (31). CNS complications should be considered in patients with compatible clinical features, and cerebrospinal fluid (CSF) analysis can aid diagnosis. In LMICs, where healthcare resources are limited, managing CNS complications is especially challenging (32). Although evidence of CNS and peripheral nervous system complications in monkeypox patients is still limited, scattered case reports highlight the need for early and aggressive treatment to prevent severe outcomes (27). The high cost of the MVA-BN monkeypox vaccine, at $100 per dose, further complicates access for people in LMICs, making them reliant on donations from high-income countries (33).

### 4.4 Cardiovascular System

There is very limited data about the cardiovascular complications of monkeypox cases in Africa which represents a greater population of the LMICs. Few cases of cardiac involvement in monkeypox have been reported, but complications such as myocarditis and pericarditis have been identified. Case reports have documented elevated cardiac biomarkers in patients with chest pain associated with monkeypox-induced myocarditis. Lymphocytic inflammation followed by myonecrosis is a common feature of viral myocarditis. Cardiac manifestations of monkeypox range from mild abnormalities to severe complications like congestive heart failure and arrhythmias (34). Symptoms such as chest pain, shortness of breath, palpitations, and edema necessitate thorough cardiovascular evaluation (35).

Viral myocarditis may be self-limiting or progress to fulminant myocarditis, which is rare (36). Diagnosis is often difficult due to the nonspecific nature of cardiac symptoms (35). Treatment requires multidisciplinary approaches, with long-term monitoring and specialized care posing challenges for LMICs, where healthcare facilities and diagnostic capabilities are limited (35).

### 4.5 Ocular Complications

The most typical ocular sign of an MPXV infection is a rash that appears in the orbital and periorbital regions (12). In a retrospective investigation, Ogoina et al. found that 25% of 40 Mpox patients treated to Nigerian hospitals between September 2017 and December 2018 had ocular rashes (12). Instead of a primary conjunctival lesion as part of a regional virus eruption, it is believed that the virus enters the eye through autoinoculation of a periocular lesion or the contiguous spread of a lesion into the eye. High-risk lesions that could put individuals at risk for eye illness include periocular or lid edge lesions. A conjunctival follicular reaction, distinct conjunctival lesions, pseudo membranes, or subconjunctival nodules are some of the different ways that conjunctival involvement can manifest. As has been previously reported in smallpox cases, there have been no instances of intraocular involvement (such as uveitis or optic neuritis). Significant scarring and bacterial superinfection can result from corneal involvement, which can manifest as stromal keratitis or corneal ulceration. This can cause either temporary or permanent vision impairment (37).

Common ocular clinical signs of MPXV infection include involvement of the eyelids and conjunctiva. Conjunctival ulcers, diffused blistering or papular conjunctival lesions, conjunctival follicular reactions, and pseudomembranous/subconjunctival nodules are some of the symptoms of conjunctivitis brought on by an MPXV infection. According to Hughes et al.’s DRC study, conjunctivitis affected 23.1% of Mpox patients in the Tshuapa health district (38).

Systemic symptoms like fever and discomfort frequently accompany these rashes and other facial abnormalities. Pink eye, also known as conjunctivitis, is mainly brought on by bacterial or viral infections in youngsters and the elderly, in part because of weakened immune systems. It manifests as an inflammatory reaction with a notable outflow of mucus from the conjunctiva. In over 20% of Mpox cases, conjunctivitis was the most frequently reported symptom. Furthermore, other symptoms such nausea, chills, sweating, mouth ulcers, sore throat, general malaise, lymphadenopathy, and photophobia were often more common in Mpox patients with conjunctivitis. Because Mpox affects both the eyelid and the conjunctiva, it can also cause blepharoconjunctivitis, or inflammation of both of these ocular regions (37,38).

The MPXV genome sequencing of an isolate from one patient identified it as belonging to lineage B1 in clade IIb. All patients had keratitis, three had uveitis (60%) and two had hypopyon (40%). Replicating MPXV was found in at least one ocular sample of all patients between day 31 and day 145 after the onset of skin lesions. The onset of ocular symptoms occurred at a mean of 21.2 days after the appearance of the first skin lesion, and they continued for an average of 61.6 days with a worsening trend until the tecovirimat treatment was initiated (37).

The fact that there have been sporadic reports of retinitis, chorioretinitis, accommodative palsy, optic neuritis, extraocular muscle palsy, suppurative dacryocystitis, retrobulbar hemorrhage with proptosis, ankyloblepharon, and lagophthalmos with exposure keratitis in previous smallpox cases may also be of interest to eye care providers, even though no reports have yet been made in Mpox patients (38,39).

### 4.6 Respiratory System

The systematic review indicates that respiratory complications associated with monkeypox (MPX) in low- and middle-income countries (LMICs) in Africa predominantly affect younger age groups. Studies from the Democratic Republic of Congo (DRC) and Nigeria— two countries reporting the highest MPX case burden in Africa—suggest that individuals aged 0–20 years are disproportionately affected by respiratory complications such as bronchopneumonia and pleural effusion (23, 40). For instance, a study in the DRC found that 65% of patients with respiratory complications were under 15 years old, highlighting the vulnerability of pediatric populations to severe MPX manifestations (40). This trend may be attributed to immature immune systems and limited access to healthcare in these regions. However, cases in older adults (40+ years) have also been documented, particularly among immunocompromised individuals or those with pre-existing respiratory conditions (23). These findings underscore the need for age-specific interventions to mitigate the burden of respiratory complications in MPX-endemic regions.

The sex distribution of respiratory complications in MPX cases from African LMICs shows a slight male predominance, though the difference is not statistically significant. Data from Nigeria and the DRC indicate that approximately 55–60% of patients with respiratory complications are male, while 40–45% are female (23, 40). This pattern aligns with broader MPX epidemiology, where males may be slightly more likely to contract the disease due to occupational exposure (e.g., hunting or animal handling) and social roles that increase contact with zoonotic reservoirs (21). However, the severity of respiratory complications does not appear to differ significantly between sexes, suggesting that environmental factors and disparities in healthcare access play a more significant role than biological differences in determining outcomes.

Respiratory complications in MPX primarily affect the lower respiratory tract, with bronchopneumonia being the most frequently reported condition. Studies from the DRC and Nigeria have identified the lungs and pleura as the most affected organs, with cases of severe bronchopneumonia leading to acute respiratory distress syndrome (ARDS) in some patients (23, 40). Although less common, pleural effusion has been documented, particularly in severe cases requiring hospitalization. Autopsy reports from the DRC have revealed diffuse alveolar damage and interstitial inflammation in patients who succumbed to MPX-related respiratory failure, highlighting the potential for severe lung pathology (40). These findings emphasize the need for early diagnosis and management to prevent progression to life-threatening complications.

The incidence and prevalence of respiratory complications in MPX cases vary across African LMICs, with higher rates reported in endemic regions such as the DRC and Nigeria. A study in the DRC found that approximately 15–20% of MPX patients developed respiratory complications, with bronchopneumonia accounting for 70% of these cases (40). In Nigeria, the prevalence of respiratory complications was slightly lower, at 10–15%, possibly due to differences in healthcare infrastructure and case reporting (23). However, underreporting remains a significant challenge, particularly in rural areas where access to diagnostic facilities is limited. These disparities highlight the need for improved surveillance and standardized reporting systems to accurately assess the burden of respiratory complications in MPX-endemic regions.

Respiratory complications are a significant contributor to MPX-related morbidity and mortality in African LMICs. In the DRC, studies have reported a case fatality rate (CFR) of 10–15% among patients with respiratory complications, with ARDS being the leading cause of death (40). Similarly, in Nigeria, the CFR for MPX patients with respiratory complications ranges from 8–12%, with higher mortality observed in pediatric and immunocompromised populations (23). Long-term morbidity, including chronic lung disease and reduced pulmonary function, has also been documented in survivors of severe MPX-related respiratory complications. These findings highlight the urgent need for targeted interventions, including vaccination campaigns and improved access to critical care, to reduce the burden of respiratory complications in MPX-endemic regions.

### 4.7 Gastrointestinal System

Monkeypox-associated gastrointestinal symptoms include anorexia, diarrhea, nausea, vomiting, and abdominal pain, with anorexia being the most prevalent (41). Hepatomegaly has also been reported as a rare complication (23). In an outbreak of monkeypox in DRC between 1996-1997, 11% of the 419 cases identified had diarrhea. Of note, 85% of the cases were under the age of 16 (42). Diarrhea and vomiting occur in the second week of illness and cause significant clinical deterioration and dehydration. These symptoms were more common in those unvaccinated against smallpox (43). Severe cases may lead to haemorrhagic colitis or gastrointestinal bleeding (44), with pediatric and immunocompromised individuals being especially vulnerable (45).

Poor sanitation in LMICs exacerbates gastrointestinal issues, increasing the burden of diarrheal diseases (25). Dehydration, a common consequence, contributes to mortality, especially in areas lacking access to fluid and electrolyte replacement therapies (25). Studies from the Democratic Republic of Congo show that patients receiving hospital care have lower mortality rates, though many arrive at the hospital already malnourished and dehydrated due to poor infrastructure (46). Recent deaths associated with haemorrhagic colitis in Nigeria emphasize the need for improved medical care in LMICs to manage monkeypox’s gastrointestinal complications (47).

### 5.0 Conclusion

Monkeypox remains a significant public health concern in African LMICs, where it has been endemic for decades. This review highlights its multisystem complications, including dermatologic, neurological, cardiovascular, musculoskeletal, gastrointestinal, ocular and respiratory manifestations, with severe outcomes in vulnerable populations. The increasing shift in cases toward young adults underscores the evolving epidemiology of the disease.

Limited healthcare access, delayed diagnostics, and inadequate treatment options worsen outcomes, necessitating urgent improvements in healthcare infrastructure, surveillance, and vaccine accessibility. Strengthening clinical management and public health preparedness is essential to mitigating monkeypox’s burden and improving patient outcomes in the most affected regions.

## Data Availability

All data produced in the present study are available upon reasonable request to the authors

## Conflict of Interest

The author declares no conflict of interest

## Funding

No funding was received for this study

## Notes

Conflict of Interest Statement: The authors declare no conflicts of interest related to this work.

Funding Statement: No funding was received for this study.

### Competing Interest Statement

The authors have declared no competing interest.

### Funding Statement

This study did not receive any funding

